# Self-attention based deep learning model for predicting the coronavirus sequences from high-throughput sequencing data

**DOI:** 10.1101/2024.08.07.24311618

**Authors:** ZhenNan Wang, Chaomei Liu

## Abstract

Transformer models have achieved excellent results in various tasks, primarily due to the self-attention mechanism. We explore using self-attention for detecting coronavirus sequences in high-throughput sequencing data, offering a novel approach for accurately identifying emerging and highly variable coronavirus strains. Coronavirus and human genome data were obtained from the Genomic Data Commons (GDC) and the National Genomics Data Center (NGDC) databases. After preprocessing, a simulated high-throughput sequencing dataset of coronavirus-infected samples was constructed. This dataset was divided into training, validation, and test datasets. The self-attention-based model was trained on the training datasets, tested on the validation and test datasets, and SARS-CoV-2 genome data were collected as an independent test datasets. The results showed that the self-attention-based model outperformed traditional bioinformatics methods in terms of performance on both the test and the independent test datasets, with a significant improvement in computation speed. The self-attention-based model can sensitively and rapidly detect coronavirus sequences from high-throughput sequencing data while exhibiting excellent generalization ability. It can accurately detect emerging and highly variable coronavirus strains, providing a new approach for identifying such viruses.

## Introduction

Infectious diseases, being the primary cause of death globally^1^, have significantly influenced the development of world history and population dynamics in different historical periods^2^. Meanwhile, with the acceleration of globalization, the movement of people and trade has facilitated the spread and evolution of infectious disease pathogens, posing significant challenges to global public health^3^. For emerging and sudden infectious diseases, rapid identification of pathogens is crucial for disease control and clinical treatment.

Coronaviruses (CoV) are a group of enveloped, single-stranded positive-sense RNA viruses that cause various diseases in mammals and birds^4^. In 2002, the Severe Acute Respiratory Syndrome Coronavirus (SARS-CoV) outbreak began in China and subsequently spread to multiple countries and regions worldwide. In 2012, the Middle East Respiratory Syndrome Coronavirus (MERS-CoV) was identified in Saudi Arabia. Globally, this pathogen caused at least 2,494 infections and 858 deaths, with a fatality rate of about 34%^5^. At the end of 2019, the novel coronavirus (SRAS-CoV-2) was first identified in Wuhan^6^. The COVID-19 pandemic is ongoing.

Quick identification of coronaviruses sequence, especially emerging and highly variable strains, is crucial for controlling and treating coronavirus outbreaks. The gold standard for coronavirus diagnosis is viral isolation followed by identification using electron microscopy in cell cultures. This method is technically demanding, time-consuming, and has low sensitivity^7^. The real-time reverse-transcription polymerase chain reaction (RT-PCR) amplification method is the preferred approach for detecting coronaviruses. This method offers real-time monitoring, high sensitivity, and specificity. However, it has some drawbacks, such as the inability to detect novel and highly variable strains, high primer requirements^8^. If a preliminary identification can be made before pathogen isolation, it can guide the isolation process, significantly improve efficiency, help quickly identify the target pathogen, determine the full genome sequence, and swiftly design PCR primers. This can buy valuable time for controlling new coronavirus outbreaks.

High-throughput sequencing, also known as next-generation sequencing (NGS), has rapidly developed over the past 15 years, becoming a widely used routine technology in the field of biomedicine. High-throughput sequencing can sequence all nucleic acids in a patient sample, and by analyzing these sequences, it can identify the pathogen present in the sample. When dealing with unknown pathogens or highly variable strains of known pathogens, high-throughput sequencing can decode the pathogen sequences without prior knowledge, guiding subsequent pathogen isolation and whole-genome sequencing. The traditional method for analyzing high-throughput sequencing data is sequence alignment. Although many sequence alignment algorithms tailored to high-throughput sequencing data, such as BWA^9^ and Bowtie^10^, have been developed, these algorithms are time-consuming and require significant computational resources.

Transformer models have achieved outstanding performance across various tasks, particularly in natural language processing. The self-attention is key mechanism of transformers models^11^. To enhance the detection efficiency and performance of novel and highly variable coronavirus sequences in high-throughput sequencing data, we designed a coronavirus sequence discrimination tool using a model based on self-attention mechanism.

## Result

### Simulated high-throughput sequencing dataset of coronavirus samples

This study selected complete coronavirus genome sequences and the human hg38 reference genome sequence to construct simulated high-throughput sequencing data of coronavirus-infected samples, which were then used for model training, validation, and testing. We obtained a total of 2,721 complete coronavirus genome sequences (excluding SARS-CoV-2), 2,779 human reference genome chromosome sequences, and 275 SARS-CoV-2 sequences **(Table 1)**.

**Table 1.** The number of coronavirus and human whole genome sequences.

| Species | Type | Source | Number of Sequences |
| --- | --- | --- | --- |
| Coronavirus<sup>#</sup> | WGS | GDC, NGDC | 2721 |
| SARS-CoV-2 | WGS | NGDC | 275 |
| Homo sapiens | WGS | GDC | 2779 |
| Total |  |  |  |

<sup>#</sup> Excluding SARS-CoV-2

After randomly dividing the coronavirus genome sequences into groups with a 80:10:10 ratio, 150 nt sequence fragments were extracted using a sliding window approach, with duplicates removed. An equal number of 150 nt fragments were randomly selected from the human genome, excluding fragments containing ‘N’. This process resulted in the final number of fragments for the training set, validation set, test set, and independent test set **(Table 2)**.

**Table 2.** Number of sequences after preprocessing.

| Data | Number of simulated sequences |
| --- | --- |
| Training | 23,188,731 |
| Validation | 2,898,591 |
| Test | 2,648,238 |
| SARS-CoV-2 | 1,812,143 |
| Total | 30,547,703 |

### Model Performance

On the simulated high-throughput sequencing dataset of coronavirus samples, each base is treated as a feature in this study. Subsequently, one-hot encoding is applied to each feature to vectorize each sequence, ensuring that the distance between each feature vector and other feature vectors is consistent and avoiding artificial errors between features. The model architecture is illustrated in Figure1. The Multi-Head attention structure is configured with five layers. Two fully connected layer is added after multi-head attention unit, which transforms the high-dimensinal output of the multi-head attention unit to a low dimentional one. Finally, the output of the linear layer is mapped to [0,1] through the Sigmoid function, with sequences judged as coronavirus sequences if the result is greater than 0.5. Additionally, the batch size for data processing is set to 1000 in this study.

**Figure 1.**
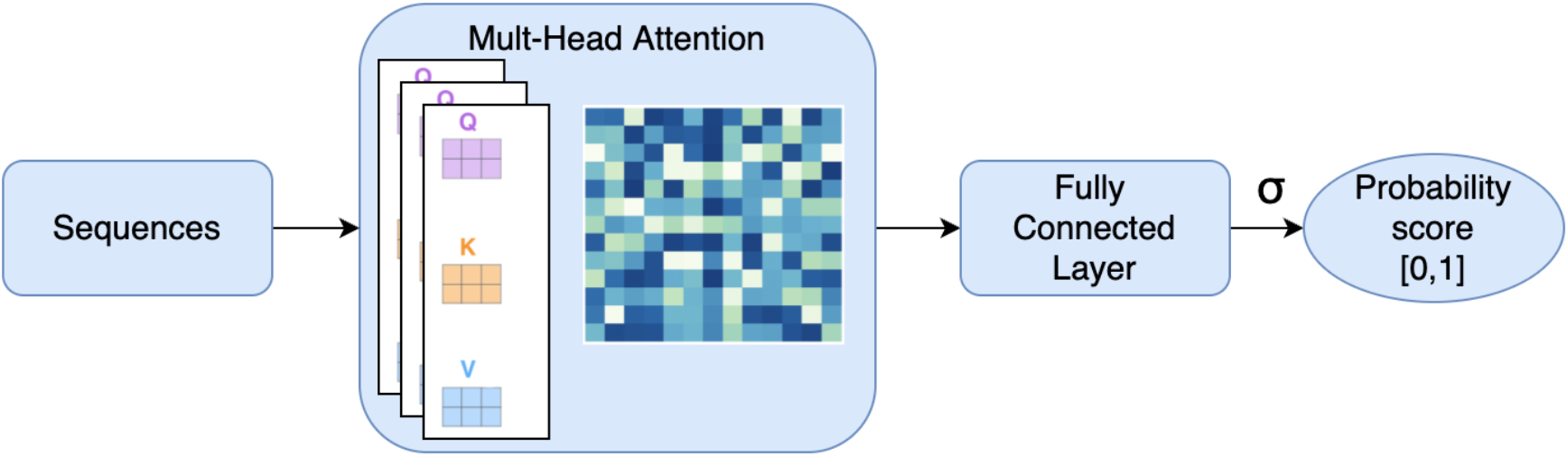
Multi-head self-attention architecture for coronavirus sequence detection.

Figure 2 illustrates the correlations among the training set accuracy, validation set accuracy, as well as the sensitivity and specificity of the validation set, in relation to the number of iterations. It can be observed that with the increase in the number of iterations, the accuracy of the training set and the validation set reached 99.33% and 98.40% respectively. The sensitivity and specificity of the validation set reached 98.22% and 99.21% respectively. Subsequently, we evaluated the model using the test set. When sequences with probability score greater than 0.5 were considered as coronavirus sequences, the accuracy, sensitivity, and specificity of the self-attention based model on the test set were 98.33 %, 98.10%, and 98.13%, respectively (see Table 3).

**Figure 2.**
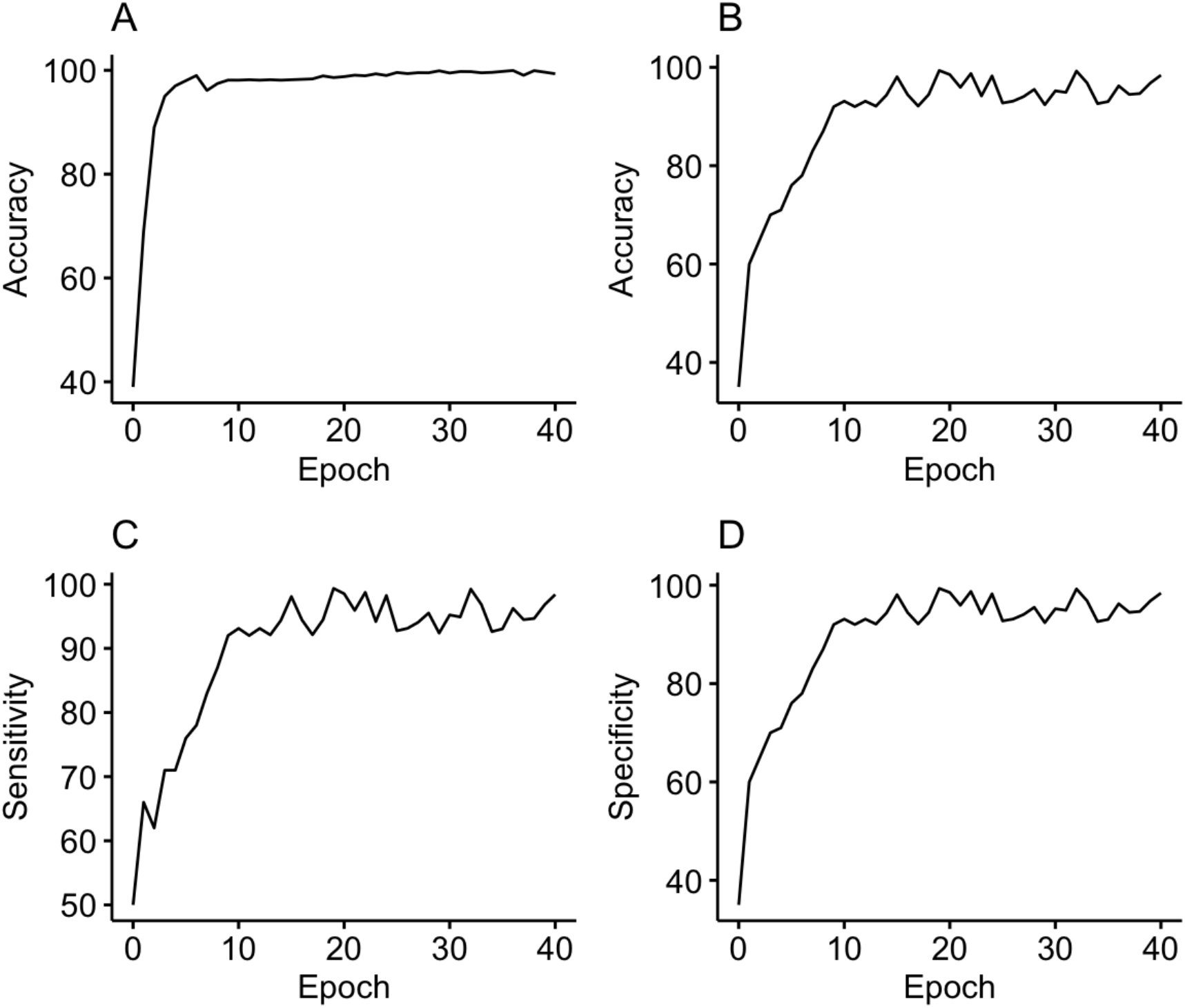
The accuracy of the training data (A) and validation data (B). The sensitivity (C), and specificity (D) of the validation set.

**Table 3.** The performance of the model.

| Data | Accuracy(%) | Sensitivity(%) | Specificity(%) |
| --- | --- | --- | --- |
| Training | 99.33 | 99.45 | 99.88 |
| Validation | 98.40 | 98.22 | 99.21 |
| Test | 98.33 | 98.10 | 98.13 |

Since a binary classification model is constructed, this study attempted to set a rejection region to avoid making “either/or” judgments on sequences. This study observed the output scores of the model for each sequence in the test set. The results showed that 98.78% of the coronavirus sequences in the test set had scores ≥0.9, while 98.98% of the human genome fragments in the test set had scores ≤0.1 (see Figure 3). Therefore, this study set the rejection region of the model to [0.2, 0.8], meaning that when the probability score is ≥0.8, the sequence is judged as a coronavirus sequence; when the probability score is ≤0.2, the sequence is judged as a human genome sequence; and when the output score is between 0.2 and 0.8, the sequence is neither a coronavirus nor a human genome sequence.

**Figure 3.**
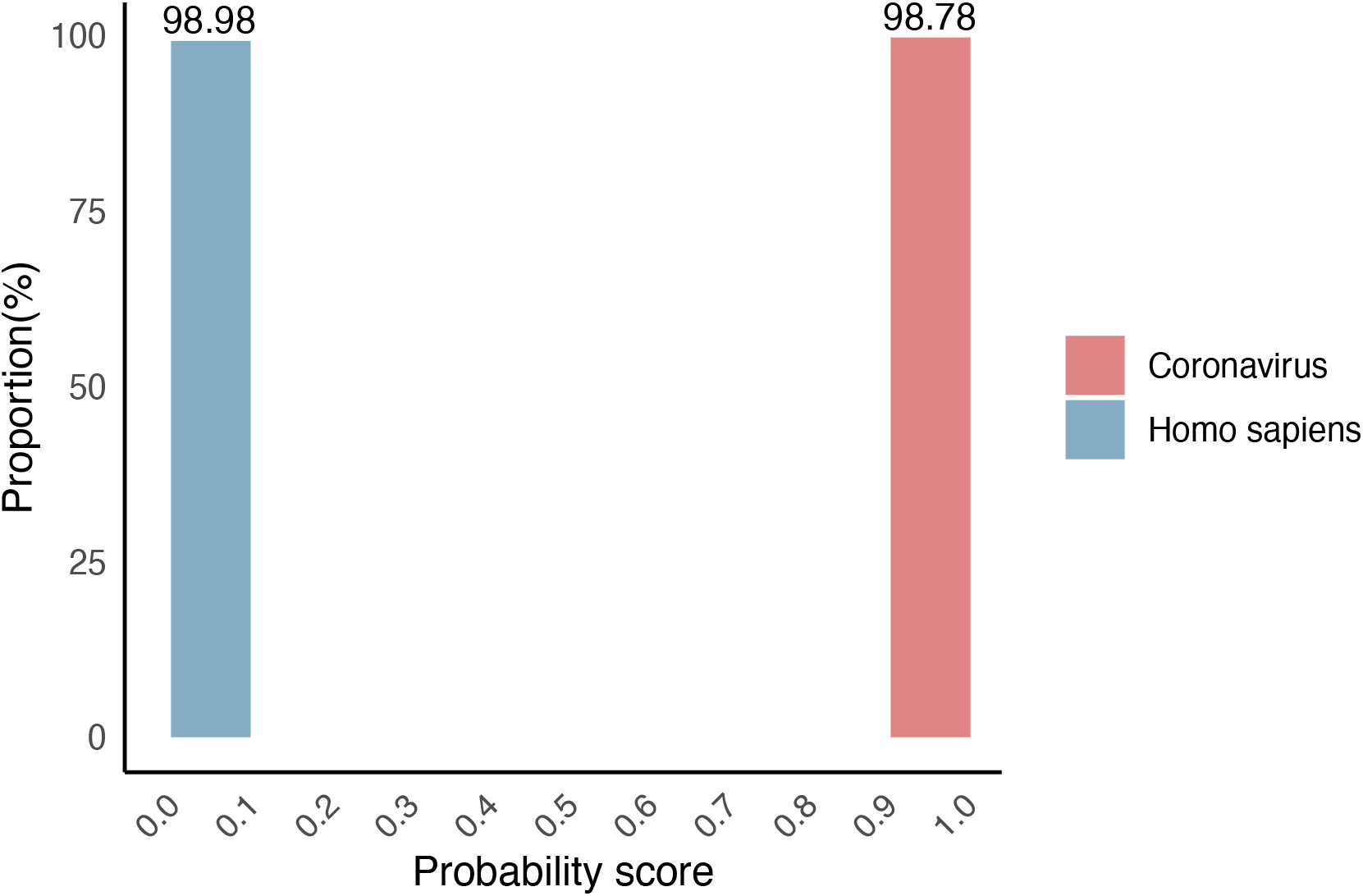
The probability score distribution of the test data.

### Comparison between the self-attention based Model and Other Classification Tools

To evaluate the performance of different tools in identifying the novel coronavirus SARS-CoV-2, we conducted a comparative analysis on an independent test dataset. The models assessed included our self-attention based model, Kraken^12^ (version 1.1.1) and Megablast^13^ (version 2.9.0). Kraken is a sequence classification software that uses k-mers for precise alignment and is capable of classifying metagenomic sequences. It is one of the most commonly used software for metagenomic sequence classification and also one of the fastest sequence classification tools available. Megablast is a classical sequence alignment algorithm known for its excellent performance and wide application in sequence alignment. For Kraken and Megablast, we constructed alignment databases using the human hg38 reference genome sequences and all coronavirus sequences excluding SARS-CoV-2. The self-attention based model used a rejection region of [0.2, 0.8], while Kraken and Megablast used default parameters, and the results of the three tools on the independent test set were shown as Figure 4. The sensitivity of the self-attention based model, Kraken, and Megablast were 97.81%, 65.80%, and 74.29%, respectively.

**Figure 4.**
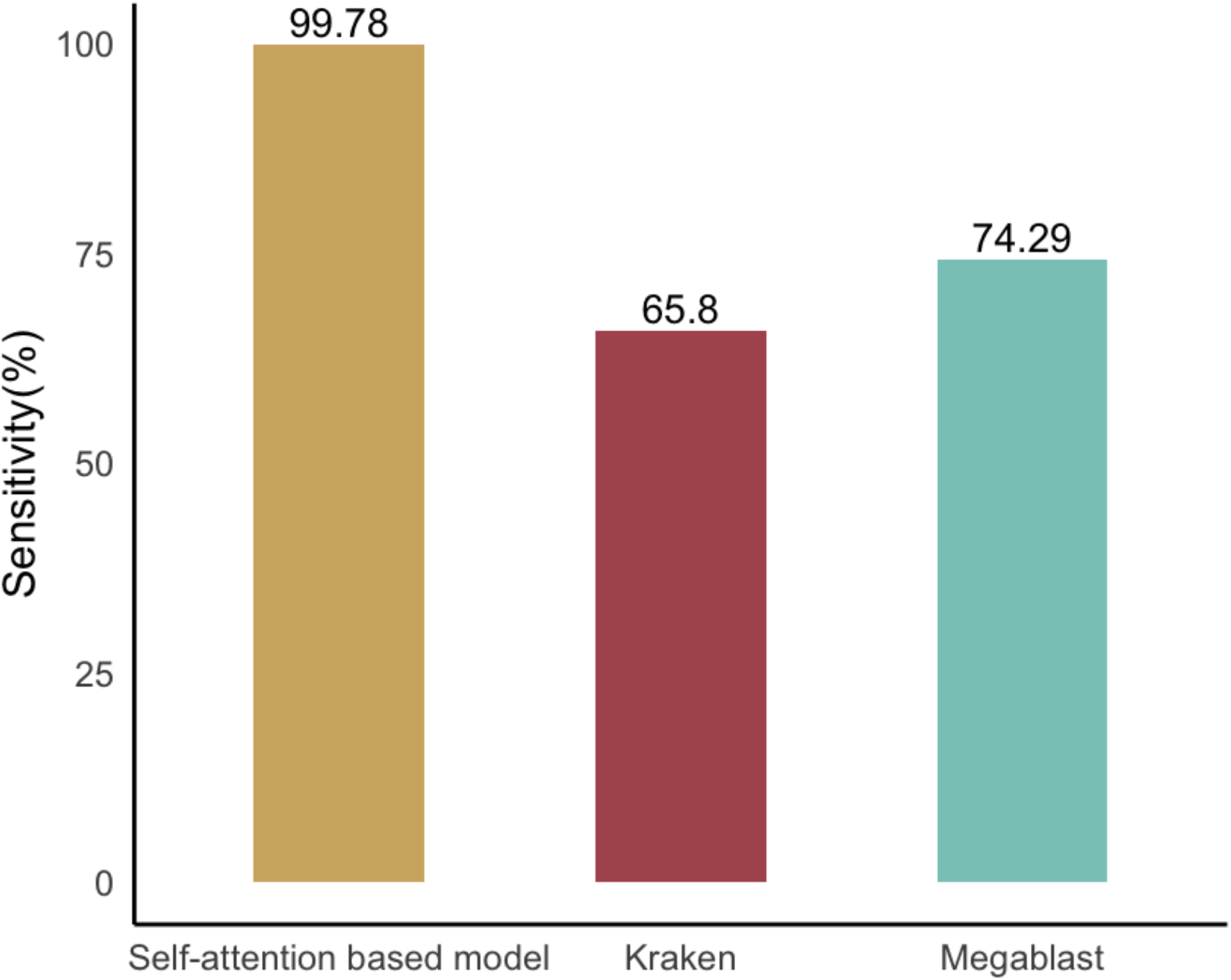
Sensitivity of the Self-attention based Model, Kraken, and Megablast on the Independent Test Set.

We compared the computational time of the model on the test set with several other common methods. The results showed that the classification speed of the self-attention based model was approximately 29% higher compared to the current metagenomic sequence classification software Kraken, and significantly outperformed Megablast (Figure 5). The computational speeds of Kraken and Megablast were obtained from the work of Wood et al^12^.

**Figure 5.**
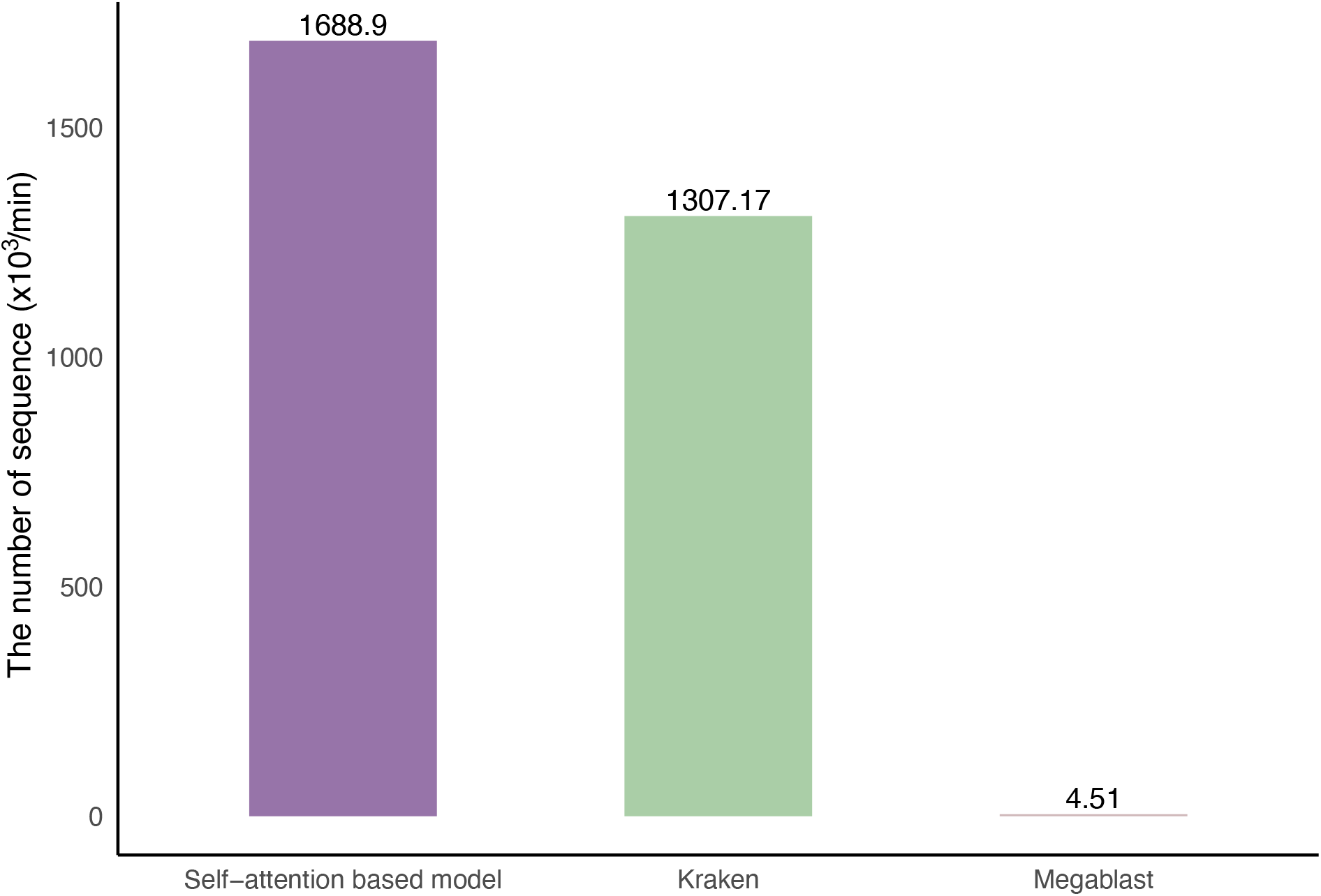
Processing speed of Self-attention based model, Kraken and Megablast.

## Discussion

In recent years, major outbreaks of emerging infectious diseases have posed challenges to the infectious disease prevention and control systems in China and globally. Rapid identification of infectious disease pathogens, especially newly emerging or highly variable strains, is crucial for the prevention and control of infectious disease outbreaks. This study collected coronavirus and human whole genome sequences, simulated high-throughput sequencing data from coronavirus-infected samples, trained a self-attention based neural network model, and developed a tool for rapid detection of newly emerging and highly variable strains of coronavirus sequences. Compared to traditional bioinformatics tools, this tool can significantly reduce computation time, lower computational resource requirements, and avoid downloading reference genomes. This tool achieved accuracy, sensitivity, and specificity of over 98% on the validation and test sets, with a sensitivity of 97.81% on the independent test dataset for SARS-CoV-2, demonstrating excellent generalization capability of the model.

The results of this study indicate that the model outperforms traditional Kraken and Megablast methods in both computational speed and discriminative performance. Once the model training is completed, the judgment of sequence categories is transformed into a single calculation between the sequence and model parameters, and the classification speed is independent of the size of the alignment database. Whereas Kraken and Megablast are fundamentally based on sequence alignment algorithms, the classification of sequences requires comparison of sequences with those in the alignment database, hence the classification speed is not only limited by the computational complexity of the sequence alignment algorithm itself, but also by the size of the alignment database. From this perspective, training the deep learning model to replace traditional sequence alignment algorithms offers significant advantages and potential in terms of computational speed.

Our work explores the application of self-attention based model for the detection of newly emerging and highly variable pathogenic agents. Although deep learning has been applied in various biomedical fields, our work represents the first attempt to apply it to the detection of newly emerging and highly variable pathogenic agents. Our research results suggest that applying deep learning to the detection of newly emerging and highly variable pathogenic agents not only yields good performance but also effectively reduces computation time and computational resource requirements. This is of great significance for the rapid detection of newly emerging infectious disease pathogens and the acquisition of valuable prevention and control time. Moreover, once the deep learning model is trained, it can be integrated into a small computing device combining software and hardware, which is easy to use and does not rely on pathogen databases, facilitating the rapid deployment of detection capabilities for newly emerging infectious disease pathogens. Therefore, conducting detection of newly emerging and highly variable pathogenic agents based on deep learning is a meaningful research direction. However, as an initial exploration, the performance of our model still needs to be tested with real samples. In the future, we will continue to expand this research, extending the range of pathogens from coronaviruses to all infectious disease pathogens, and from binary classification models to multi-classification models, to enable better performance in practical applications.

## Methods

### Data collection

In this study, we downloaded all available full-genome sequences of coronaviruses from the NCBI database. As of December 16, 2023, we collected a total of 2,721 sequences. Additionally, we obtained the human reference genome sequence from the NCBI Genomic Data Commons (GDC) and the SARS-CoV-2 WGS from the National Genomics Data Center (NGDC). As of February 19, 2023, we acquired 275 sequences.

### Data Preprocessing

First, we divided the obtained full-genome sequences of coronaviruses into training, validation, and test sets in a ratio of 80:10:10. Given that current high-throughput sequencing typically uses PE150 sequencing with a read length of 150 nt, we set the sequence length for simulated sequencing data to 150 nt to ensure the model is applicable to PE150 high-throughput sequencing data. We employed a sliding window with a length of 150 nt and a step size of 1 nt to extract continuous sequences of 150 nt from the training, validation, and test sets of the coronavirus genomes. Duplicate sequences within each dataset were then removed.

Considering that the length of the human hg38 reference genome far exceeds the length of coronavirus genome, we used an equivalent length window to randomly select 150 nt fragments from the human hg38 reference genome to balance the data volume between the two types of datasets. Consequently, the number of fragments from the human reference genome matched those of the coronavirus training, validation, and test sets.

Since genomic sequences contain not only the four bases (“A”, “T”, “C”, and “G”), but also “N” to denote ambiguous bases, we discarded any fragments containing “N” during data preprocessing. For the SARS-CoV-2 sequences, we applied the same preprocessing rules and used these sequences as an additional independent test set.

### Calculation of Accuracy, Sensitivity, and Specificity

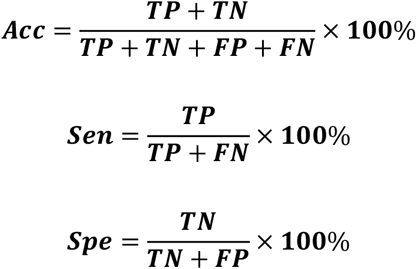

### Computing environment

CPU: Intel Core i5-10400

RAM: 128 GBytes DDR4 2666 MHz

GPU: NVIDIA RTX 3090

OS: Ubuntu 20.04

Python: 3.8.10

Pytorch: 1.81

CUDA: 11.2

## Data availability

All the genome data in this study are obtained from public datasets which available in the GDC or NGDC database.

